# New Insights into Batten Disease CLN1 from a Patient Facing Registry

**DOI:** 10.1101/2025.09.25.25336690

**Authors:** Sean Ekins, Scott H. Snyder, Melanie Tojong, Thomas R. Lane, Meena Hamilton, Emily C. de los Reyes

## Abstract

There is currently no FDA approved treatment for Batten disease CLN1, a rapidly progressive, fatal, and rare pediatric neurodegenerative disease. In this article, we aimed to collect retrospective clinical data from the families to identify CLN1 patients for a future patient facing natural history study and clinical trial. We have therefore developed and launched an institutional review board approved Batten disease CLN1 registry (https://cln1registry.collaborationspharma.com). We implemented a range of clinically relevant questions for the caregiver to answer in the registry created using the Django 5.0 web framework. Authenticated users provide profile data as well as patient and caregiver information, which is collected, stored in an encrypted SQLite database and deidentified before export and analysis. The registry application is deployed on an in-house private server using Docker with the database file and decryption key stored outside of the docker image and managed independently. We analyzed the collected data from 23 individuals with CLN1 Batten disease, including geographic distribution, age of diagnosis, onset of disease including motor and language declines. Although CLN1 is well known as infantile onset (<18 months), on average the diagnosis was not identified until 25 months which indicates a delay in diagnosis. We identified how the onset of disease is similar with most occurring before 25 months while infantile and late infantile patients had earlier onset and more frequent seizures in comparison to juvenile onset patients. Antiepileptic medications were the most heavily represented. Our findings provide new insights for CLN1 clinical care, enabling readiness for future natural history and clinical trials.

## Introduction

The neuronal ceroid-lipofuscinoses (NCLs) are lysosomal storage diseases (LSDs) and collectively are the most common hereditary progressive neuro-visceral degenerative disease of children with a prevalence of approximately 1.5 to nine per million population (1.3 to 7 per 100,000 live births) (Mole and Williams, 1993). Deficiency of the lysosomal enzyme palmitoyl protein thioesterase-1 (PPT1; EC 3.1.2.22) causes a human lysosomal storage disorder, (CLN1 disease). CLN1 is characterized by progressive blindness, motor and language regression, seizures, ataxia, and early death (Kohlschütter et al., 2019) The onset of the disorder varies from infancy to adulthood (Das et al., 1998; Ramadan et al., 2007; van Diggelen et al., 2001). Currently over 90 different biallelic mutations of PPT1 have been reported in CLN1 patients (Augustine et al., 2021; Kousi et al., 2012; Mole and Cotman, 2015/10/01). One earlier study included 29 different patients with mutations (Hofmann et al., 1999) in the US (Kohlschütter et al., 2019). To date there are no treatment options available for this fatal disorder (Masten et al., 2020b) except for symptom management of seizures, movement disorders, sleep, pain, mood and palliative care (Augustine et al., 2021). There have been some efforts towards gene therapies, small molecule treatments and other biologics such as enzyme replacement therapy (ERT) (Hahn et al., 2022; Johnson et al., 2019; Masten et al., 2020b). There have also been very limited clinical trials for treatments for CLN1. One of these clinical trials evaluated cysteamine and *N*-acetylcysteine. This latter study included electroencephalography (EEG) and neuroimaging with magnetic resonance imaging (MRI) and (MRS) spectroscopy analysis. The treatment did not improve the neurodegenerative course of the disease even though it decreased a granular osmiophilic deposits in leukocytes (Levin et al., 2014). It has also been demonstrated in preclinical studies that a single dose of purified recombinant human PPT1 (rhPPT1) delivered intravenously as an ERT to neonatal *Ppt1^−/−^* mice corrected the enzyme defect, and potentially corrective levels were observed in heart, lung and spleen in mouse (Hu et al., 2012). In a subsequent study, intrathecal delivery of a single dose of PPT1 (Lu et al., 2015) produced moderate improvements in motor function and survival. Most recently, monthly delivery of rhPPT1 by intracerebroventricular (ICV) dosing statistically significantly altered the trajectory of a knockout mouse model of the disease as assessed by various phenotypic and histologic assessments (Nelvagal et al., 2022).

### Treatments for Batten disease

These efforts towards an ERT are following on the FDA approval of Cerliponase alfa (Brineura; BioMarin Pharmaceutical) (tripeptidyl peptidase 1, rhTPP1), delivered via ICV infusions to treat CLN2 Batten disease. Safety and efficacy were evaluated in patients with CLN2 Batten in a Phase I/II study (NCT01907087). Efficacy of the ERT in patients was monitored with the CLN2 rating scales for motor–language scores and total scores in four domains namely, motor skills, language, vision and seizures (Schulz et al., 2018) and compared with natural history data from 42 patients from two CLN2 Batten disease registries (Weill Cornell CNS (Worgall et al., 2007) and Hamburg CLN2 scale (Steinfeld et al., 2002)). Rates of decline from baseline in the motor–language score were calculated over a primary 48-week period (Schulz et al., 2018) and a 240 week extension study (Schulz et al., 2024). Treatment with Cerliponase alfa was significantly less likely than historical untreated controls to have an unreversed two point decline indicating a slowing of motor and language disease progression in children with CLN2 Batten disease (Schulz et al., 2024). It is hypothesized that rhPPT1 when delivered ICV would have a similar dramatic effect for CLN1. The FDA has issued Collaborations Pharmaceuticals, Inc. (CPI) an orphan drug designation (#16-5701) and a rare pediatric disease designation for developing rhPPT1 (CPI-601, #RPD-2020-302) (Nelvagal et al., 2022).

### Registries for rare diseases

Regulators have recognized that real world evidence (RWE) is important for drug approvals and it can be used as controls in clinical studies as well as assist in patient recruitment (Boulanger et al., 2020; FDA, 2018). Patient and public involvement in real world data through registries is one way to inform researchers and healthcare providers about disease progression and interventions. A patient registry documents and stores detailed information about a specific disease or syndrome and the characteristics can vary from patient-, disease-based and product-based registry (Pisa et al., 2023). Guidance is available from the FDA, EMA (Agency, 2023) and others (Hageman et al., 2023; Kolker et al., 2022; Mordenti et al., 2022; Wicks et al., 2023) on the building and use of registries for supporting regulatory decision making for drugs and biological products. Subsequently, many rare diseases have one or more registry (clinic entered data or patient/caregiver (patient facing) entered data) (Hageman et al., 2023) e.g. Gaucher disease (Collin-Histed et al., 2023), adult polyglucosan body disease (Sparks et al., 2024), Muscular dystrophy (Kilroy et al., 2023), Charcot Marie Tooth (Sames et al., 2014) that have gone on to identify unmet needs in disease management. All the registries developed are unique (Boulanger et al., 2020) as there is not a single universal model. Some rare disorders such as Gaucher disease have multiple registries including proprietary ones from pharmaceutical companies as well as those from individual countries (Elstein et al., 2024).

### Natural history studies for Batten disease

In general, collection, analysis and dissemination of data on disease progression and patient responses to long-term disease management strategies represents an important way to improve understanding of the disease and keep medical professionals up to date on the latest advances as well as provide a natural history of a condition (Wicks et al., 2023). This is particularly important for rare diseases were there are very few patients. A natural history study (NHS) is valuable in understanding disease progression and more (Liu et al., 2022). In rare diseases, NHS have been critical in cases where randomizing subjects to a placebo arm may not be feasible or ethical due to the limited number of patients (Li and Izem, 2022). Retrospective NHS data have often been more valuable than that from randomized trials because of the restricted number of patient numbers (Benjamin, 2017). NHS have also benefitted patients with rare diseases by making them aware of the current standard-of-care practices and in identifying rare disease centers (FDA, 2019). Patients included in NHS become historical controls for studies without an internal control, allowing the effectiveness of the treatment to be determined (Jahanshahi et al., 2021). Natural history data therefore serves as control data in clinical trials to shorten time in drug development. The FDA (FDA, 2019, 2023) and EMA (EMA, 2020), have released guidelines to ensure high quality of natural history data for drug development in rare diseases. Published natural history data does not currently exist for CLN1 unlike for CLN2 (Nickel et al., 2018), CLN3 (Masten et al., 2020c), CLN5 (Simonati et al., 2017) and CLN6 (O’Neal et al., 2024). Disease staging for CLN1 only exists for the infantile form (Vanhanen et al., 2004) hence in preparation for a future NHS and clinical trial we need to identify CLN1 patients.

### The need for a registry for CLN1

While multiple physician-led registries have included CLN1, this is the first dedicated patient registry for CLN1 Batten disease. An early Batten disease registry was the European Concerted action NCL clinical case registry (Williams et al., 1999) which captured data on 60 patients across different types of the disease but did not provide any data analysis. An initial CLN1 natural history study from the University of Rochester was reported as an abstract (Masten et al., 2020a) which collected data for 20 CLN1 patients and disease progression was scored with the Unified Batten Disease Rating Scale (UBDRS), while their analysis suggested variation in age of onset and type of symptoms. As none of this registry data is publicly available to us (or others), we have developed a new registry to connect to current CLN1 patient families, and provide information for other healthcare providers, researchers, clinical investigators, regulators and policy-makers. The Batten Disease CLN1 registry is currently being used to collect data on patients who have either been clinically diagnosed by a doctor or genetically diagnosed with CLN1. Due to the small and heterogenous nature of population in a rare disorder like CLN1, there are many challenges associated with patient identification and recruitment which limits the statistical power of future clinical studies.

## Methods

### Participants

Participants provided consent to share deidentified data. This study was approved through a local Institutional review board (IRB) approval: WCG IRB Protocol #20241428.

### Registry Software Application Details

The CLN1 Batten registry (Ekins) was created using the Django 5.0 (Anon, 2024b) web framework. Django is an open-source web framework based on the Python programming language. It was designed to provide a complete implementation of the model/template/view architecture for deploying complex websites with built in integration with many types of databases. It supports authenticating users through a forms-based login as well as a built-in object relational mapping library (Django ORM) that provides transparent storage of software artifacts to an external database.

#### Data storage

User information as well as profile data, is collected and stored in an SQLite (Anon, SQLite) database containing contact information for users after they register as well as a profile that contains patient as well as caregiver information. SQLite is a software library that is embedded in most application development environments and is distributed with the Python runtime environment. Data in SQLite is stored in a single file although it is accessed through standard SQL (Structured Query Language) queries and appears as a set of relational database tables. All data is stored encrypted when written using a django library django-cryptography (Anon, 2024c) and decrypted when read on the website. This makes it impossible for anyone accessing the SQLite database file from reading the data outside of the website. Because we anticipate the total number of users of this registry to be no more than a few hundred, the use of SQLite as an embedded database engine simplified the system design and allowed us to deploy the database without the need for a separate database server.

#### Data retrieval

We have installed django-admin-csvexport (Anon) a library that adds CSV export functionality to the standard Django administration panel. The registry administrator can retrieve user contact information to provide further information to users that log into the site as well as retrieve anonymized profile data for further analysis.

#### Web page design

The displayed web pages use a CSS (Cascading Style Sheet) library called Bootstrap 4 (Anon, 2024a) that provides the visual representation of the web site. Programmatic form generation was performed with django-crispy-forms (Anon) which maps form elements to specific classes with predefined style information. The save function was implemented whenever the user clicks “Save” or toggles to a different section of the form. A function was added that warns the user if they have any unsaved changes when they try closing the page.

#### Application deployment

The registry application (Ekins) is deployed on our server using Docker (Anon) a containerized deployment library which allows us to create application images that contain the registry application and all of its dependencies. These images are deployed on our servers but can be redeployed on other servers as necessary. The database file and decryption key information are stored outside of the Docker image and managed independently of the application.

### Statistical analysis

For those families that provided informed consent, their deidentified registry data was downloaded (Jan 2025) and where necessary data was cleaned and obvious typographical errors corrected. For developmental milestones, a zero was considered when the child did not meet the specific milestone and was subsequently removed from statistical analysis. In addition, drug names were cleaned to deconvolute synonyms to a single drug name. Summary statistical analysis and graphing was generated using several software tools (Python vers 3.12, Matplotlib, Pandas, Seaborn, Plotly, Scipy and scikit-posthocs packages as well as Microsoft Excel).

## Results

We developed and launched the IRB approved Batten disease CLN1 registry (Ekins) on April 11^th^, 2024, (Figure 1, Figure S1) followed by a press release and sharing of the URL with CLN1 families. This registry consisted of basic contact information, clinical and caregiver information as well as permissions (Figure S1). Based on user feedback the registry also underwent a redesign in late 2024 to improve clarity for users and ensure the data input was automatically saved. This was communicated to families in December 2024. To date we have obtained data on 23 patients (Figure 1) (12 male, 11 female (Figure S2A) – 18 Caucasian, 1 Asian, 1 black, 2 mixed and 1 other, Figure S2B) whose parents/ caregivers provided informed consent to include their deidentified data in this analysis. Most of the CLN1 patients who submitted data were from the United States (US) (Figure 1).

**Figure 1.**
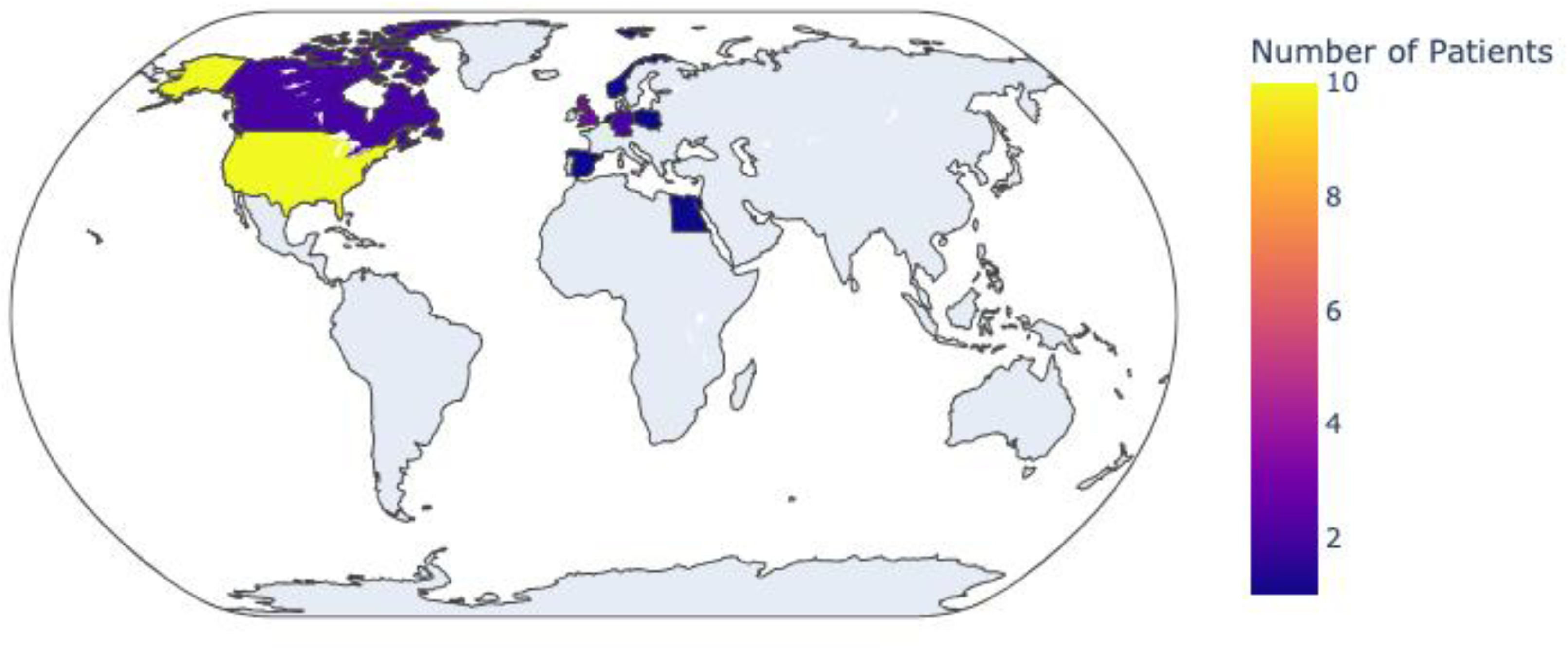
Distribution of CLN1 patients from the Registry.

Summary mean data are shown for the questions with numeric values (Figure 2, Table S1) in which patients grouped by the classifications we used to represent infantile, late infantile and juvenile CLN1 (Augustine et al., 2021). We have used the following definitions for Infantile (diagnosis at <18 months), Late Infantile (diagnosis at 18 months to less than 4 yrs) and Juvenile (diagnosis between 4 yrs and 18 yrs). Most of the patients (N = 14) had a late infantile onset (based on these definitions).

**Figure 2.**
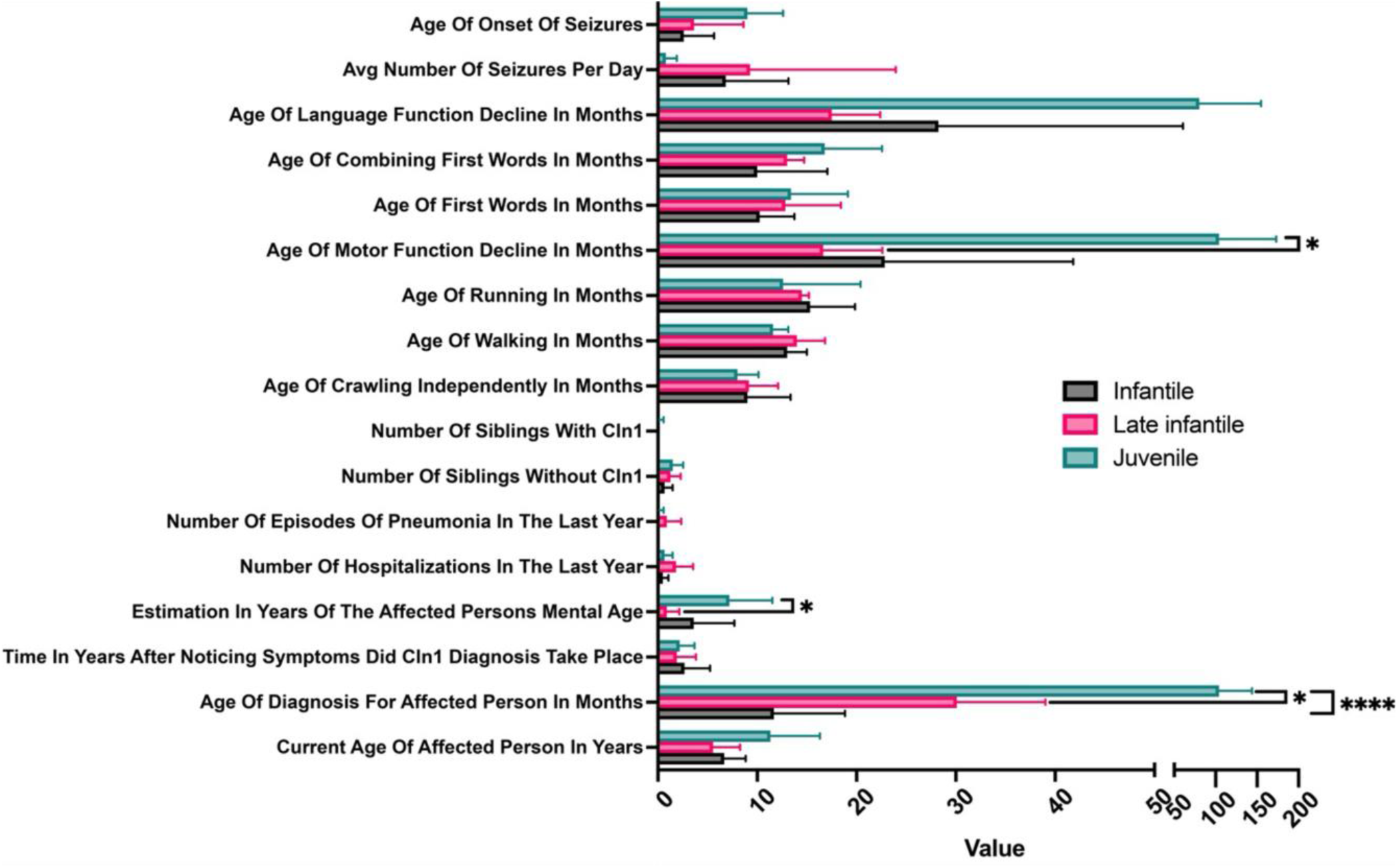
Analysis of patient data from the CLN1 registry separated by class (based on age of diagnosis). p-value significance <0.5-0.0332*, <0.0001****.

In addition, we have provided distribution plots with smooth average curve fit of the data (Figure S2). The mean age of diagnosis (Figure S2C), age of motor function decline (Figure S2D) and language function decline (Figure S2E) also track with these classifications (Table S1). Motor and language function declines are very similar with most occurring before 25 months of age in both infantile and late infantile, while these declines are much later in juvenile patients (Table S1). The first clinical sign noted by the parents was predominantly motor abnormalities, followed by seizures (Figure S2F). This concurred with age of onset of seizures (Figure S2G). The mean estimated mental age (Table S2, Figure S2H) showed a distribution lower than the current age of the patients (Table S1, Figure S2I). Late infantile patients had more hospitalizations (Fig. S2J) and episodes of pneumonia (Table S2, Figure S2K) which also followed the higher average number of seizures per day (Table S1). Juvenile patients have a markedly later age of onset of seizures (Table S1). Families reported that the inability to meet key developmental milestones (Figure S2L-P) is one of the clues that a child may have a disorder which leads families on their diagnostic odyssey (Table S3). The age of both motor function and language function decline is earlier in late infantile than infantile, however, there is also a large standard deviation in the infantile data (Table S1). The age for those children that were able to meet language and motor milestones was broadly similar across groups (Table S3). One of the patients had a sibling with CLN1 (Figure S2Q) although several had unaffected siblings (Figure S2R).

Infantile and late infantile patients have a higher average number of seizures/day and an earlier onset than in juvenile patients (Table S1, Figure S2S). The most common type of seizure was myoclonic (Figure S2T). Time after noticing symptoms to CLN1 diagnosis (Figure S2U) was approximately 2 years on average across all classes of CLN1 patient (Table S2).

The data distributions for each category were not found to pass normality tests (D’Agostino & Pearson, Anderson-Darling, Shapiro-Wilk and Kolmogorov-Smirnov tests), so these data were analyzed using a non-parametric approach. In short, all three classes (Infantile, Late Infantile, Juvenile) were assessed for significant differences at the categorical level (Kruskal-Wallis) and when statistically significant differences were found between groups a follow-up test (Post-Hoc Dunn’s test) was performed. Statistically significant differences were found between all classes with “age of diagnosis”. There were also statistically significant differences in “the age of motor function decline” and the “mental age” between the Juvenile and Late Infantile classes (Figure 2, Table S4).

Analysis of the medications reported by the CLN1 families suggests multiple anti-seizure drugs (Figure S2V) including valproic acid, levetiracetam, clonazepam, lacosemide, zonisamide, rufinamide, cannabidiol and carbamazepine were used. The most frequently reported medication is valproic acid followed by levetiracetam. This emphasizes the intractable nature of the epilepsy seen in children with CLN1 disease (Figure S2W). Another interesting observation when we further segment the first clinical sign data is that for late infantile, most children are identified by their motor abnormalities (Figure S2X).

Limited caregiver information was also obtained (when provided) including: gender (19 female, 2 male); race (20 Caucasian, 1 Asian); relationship (all parents); employment status (12 employed, 9 unemployed); marital status (19 married, 2 single) and education level (2 graduate school, 15 bachelors level, 2 college no degree, 2 high school education).

## Discussion

In order to perform a future NHS, we needed to establish the global prevalence of CLN1 disease henceforth, we required an updated registry for this disorder because of the lack of available data in the public domain. While there are numerous publications on the development of rare disease registries there are no well-defined templates. We chose elements from prior publications and had discussions with experts and families to learn from other rare disease groups (Anon, 2025; Sames et al., 2014) to develop the registry for CLN1 which captures basic patient and caregiver information. With a patient-facing registry we are required to develop a user-friendly interface so parents/ caregivers can conveniently upload their basic information with minimal guidance. These aspects of the software design benefited from our in-house expertise and feedback from CLN1 families. The company had little ‘human subjects research’ experience prior to this and had to undergo mandatory training before submission of the IRB application. This is an important point to note for others that might be considering building and running a registry for the first time.

Our findings from this registry suggest that most of the CLN1 patients are from the USA and are diagnosed early on, but it is also taking ∼2 yrs or more for a diagnosis. Although CLN1 was designated as infantile onset, this registry discovered that the children were generally not diagnosed until after 18 months of age. Age of onset, frequency, and semiology of the seizure types were also reported by the families. Seizure control and types of seizures described indicates that prevention and treatment are considered a priority to the families. Other concerns include motor and language function decline. For example, ERT treated CLN2 children declined over time in comparison to natural history controls (Schulz et al., 2024). Some children also do not meet any expected developmental milestones like independent ambulation which makes it more difficult to interpret the data collected here. This lack of acquisition of independent walking also indicates a much earlier onset of disease and the need to identify an underlying diagnosis if a treatment becomes available. It is also clear that some children have episodes of pneumonia (Table S2) and GI issues, and these may result in the use of steroids and therapies to aid digestion, respectively (Figure 2). We identified the extensive use of antiepileptic medications as well as many other classes of drugs to address various symptoms e.g. for sedation, muscle spasticity, GI issues etc. (Figure 2, Figure S2V). In one case, a patient had been treated with a gene therapy for CLN1 (Figure S2V).

### Methodological considerations and limitations of this study

Disease specific clinical rating scales have been developed for infantile, late-infantile and juvenile phenotypes of Batten disease and can be used for prospective, retrospective or combined analysis for rating the severity and progression of key symptoms of the disease. The most common scales used for monitoring Batten disease progression are the UBDRS developed by the University of Rochester (Marshall et al., 2005) and the Hamburg LINCL scale developed by DEM-CHILD (Steinfeld et al., 2002) in Europe. Other developmental scales that can be used include the Bayley Scale of Infant and Toddler development and new scoring systems which use a customized rating system to suit the disease and patient population of interest (e.g. CLN5 Batten). The data from this registry may highlight aspects that we need to capture in future scales. An initial CLN1 natural history study (Masten et al., 2020a) with data on 20 CLN1 patients was scored with the UBDRS. The results from this registry are consistent with previous reports that motor decline appears to be the most common first clinical sign observed (Figure S2F).

Limitations of this study include the limited sample size and geographic distribution of the patients. This could likely be improved by increased visibility of the registry, and this report should help in that respect. Integration with other registries might increase the size of the database although we would have to check for any patient overlap. The retrospective nature of the data collection including ascertainment of the precise date for the loss or lack of acquisition of milestones is also a limitation. We also relied on the families or caregiver input of the data versus a health professional, and this resulted in several typographical errors in drug names and the need for corrections from brand names to generic names as well as data deconvolution to address these synonyms. We also grouped patients using classifications and age cut-offs for infantile, late infantile and juvenile CLN1. In this study, we used the age of diagnosis to classify patients into the groups (Augustine et al., 2021) rather than ‘age of onset’. Other registries like DEM-CHILD may use different definitions for these groups (Nickel and Schulz, 2022; Williams and Mole, 2012). It is likely with the average time to diagnosis of ∼2 years that there may be some overlap. We can be confident in the assignment of the infantile class but some in the late infantile class may overlap closely. There were also no adults with data in our registry, so our analysis is silent in this respect. In addition, there were more late infantile patients in this analysis than the other two groups combined. These skewed, relatively small group sizes hamper statistical analysis and subsequently only a few statistically significant differences were identified (Figure 2, Table S4). Clearly this registry did not include clinical rating scales and was not intended to capture longitudinal data which would occur in a future natural history study.

Establishing the Batten disease CLN1 registry may address three critical needs. First, those who study CLN1 will need accurate information to understand the disease compared to other CLN. This study underscores the delay in diagnosis for this population which exhibits an early delay in acquisition of milestones, specifically walking. Secondly, a more formal natural history study needs to be undertaken by health care experts to identify the precise staging of vision, language and motor loss. It may also be helpful to identify the exact semiology of seizure types and classification in children with CLN1 disease since this has not been formally studied in a structured fashion. A formal natural history will also allow researchers and physicians to access patients for physical samples like blood and urine to advance research for CLN1 biomarker development. Additionally, clinicians, pharmaceutical and biotechnology companies, which are ready to begin trials, would need to communicate upcoming NHS and clinical trial details and development of the registry enables this.

### Conclusion

Our efforts to develop treatments for CLN1 will ultimately bring undiagnosed patients to the fore due to the increased visibility and awareness. Advances in ICV administration of enzyme and gene therapies has improved the chances of finding treatment options for children with CLN1 disease. There is a need to prepare for the NHS that is critical in allowing for comparisons for any clinical trial. The Batten disease CLN1 registry will help us to continue to learn from these patients and inspire others that such databases can be built and are of value in the process of developing a new treatment. It is also possible that the data collected may aid regulatory submission as it provides a snapshot of the CLN1 population and the effects the disease has on them. Could future treatment of the disease allow the patient to meet their developmental milestones, increase their mental age, decrease the incidence and frequency of seizures, delay the age of onset of seizures and decrease the number of drugs for seizures, as well as decrease the number of hospitalizations in a year or even decrease the number of drugs used? These are just some of the many questions to ask of a future treatment in a clinical trial that are derived from this study and the CLN1 registry may effectively provide a baseline for comparison.

## Data availability

All deidentified data are available from the corresponding author upon written request.

## Software availability

The registry software developed is proprietary and available to license from Collaborations Pharmaceuticals, Inc.

## Acknowledgements

The authors kindly acknowledge all the CLN1 families that have participated in the registry, and we sincerely thank Mrs. Jennifer Palermo, who has helped raise awareness of these efforts. We are also grateful for the discussions and assistance from Ms. Sarah Negri and Dr. Barbara Kroner (RTI) as well as registry logo design by Ms. Stefanie Andersen. Further discussions and considerable support from Dr. Renuka Raman, Dr. Morgan Barnes and our colleagues are also greatly appreciated.

## Author contributions

SE: conceptualization, data curation, formal analysis, funding acquisition, investigation, project administration, resources, supervision, roles/writing - original draft, and writing - review & editing; SHS: formal analysis, software, visualization, roles/writing - original draft; MT: software; TRL: formal analysis, visualization, roles/writing - original draft; MH: formal analysis, visualization; ECdlR: formal analysis, investigation, methodology, writing - review & editing.

## Financial support

This work was funded by Collaborations Pharmaceuticals, Inc.

## Conflict of interest statement

Sean Ekins is owner of Collaborations Pharmaceuticals, Inc.

Scott H. Snyder is an employee of Collaborations Pharmaceuticals, Inc.

Melanie Tojong is an employee of Collaborations Pharmaceuticals, Inc.

Thomas R. Lane is an employee of Collaborations Pharmaceuticals, Inc.

Meena Hamilton is an employee of Collaborations Pharmaceuticals, Inc.

Emily C. de los Reyes has no competing interests.

## Supplemental Information

The following additional material may be found online

**Figure S1:** The Batten Disease CLN1 registry questions on each form.

**Figure S2.** Distribution plots from the deidentified data analyzed from the CLN1 registry with smooth average curve fit of the data. A. Gender of patient, B. Race of the affected person. C. Age of diagnosis for affected person, D. Age of motor function decline, E. Age of language function decline, F. First Clinical Sign, G. Age of onset of seizures, H. Estimation in years of the affected persons mental age, I. Current age of affected person, J. Number of hospitalizations in the last year, K. number of episodes of pneumonia in the last year, L. Age of first words, M. Age of combining first words, N. Age of crawling independently, O. Age of walking, P. Age of running, Q. Number of siblings with CLN1. R. Number of siblings without CLN1, S. Average number of seizures per day, T. Seizure type, U. Time in years after noticing symptoms did CLN1 diagnosis take place, V. Drug classes used to treat symptoms in the CLN1 patient registry. W. Drugs used to treat symptoms in the CLN1 patient registry. X. First clinical sign grouped by disease classification.

**Supplemental Table 1.** Summary data for questions from Batten Disease CLN1 Registry.

**Supplemental Table 2.** Summary data for questions from Batten Disease CLN1 Registry.

**Supplemental Table 3.** Summary data for questions from Batten Disease CLN1 Registry.

**Supplemental Table 4.** Table of p-values to assess statistical significance in the Post-Hoc Dunn’s follow-up test.

## Notes

### Funding Statement

This study was funded by Collaborations Pharmaceuticals, Inc.

### Author Declarations

IRB of WCG (IRB Protocol #20241428) gave ethical approval for this work

