## Supplemental files for "New Insights into Batten Disease CLN1 from a Patient Facing Registry"

**Figure S1.** The Batten Disease CLN1 registry questions on each form.

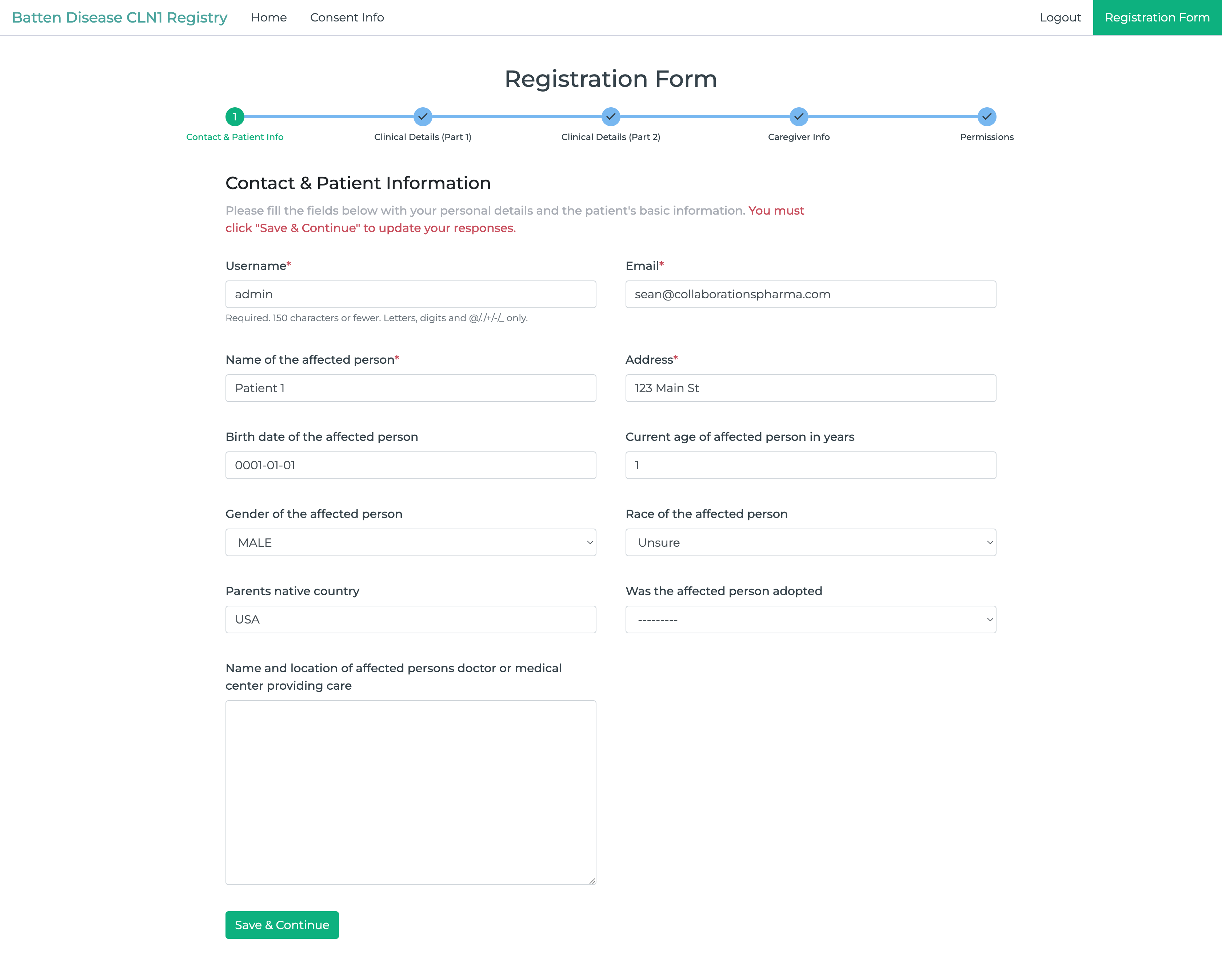

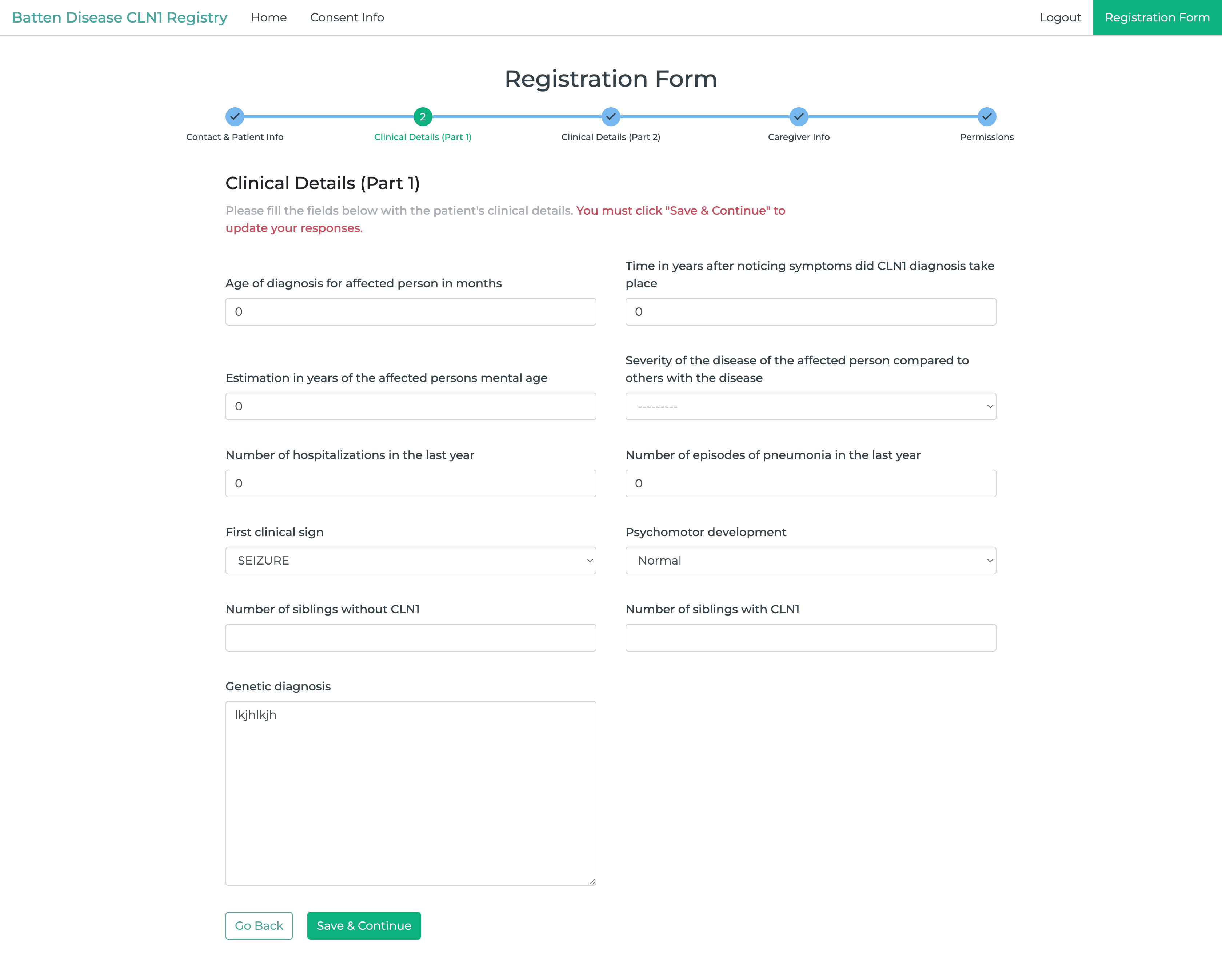

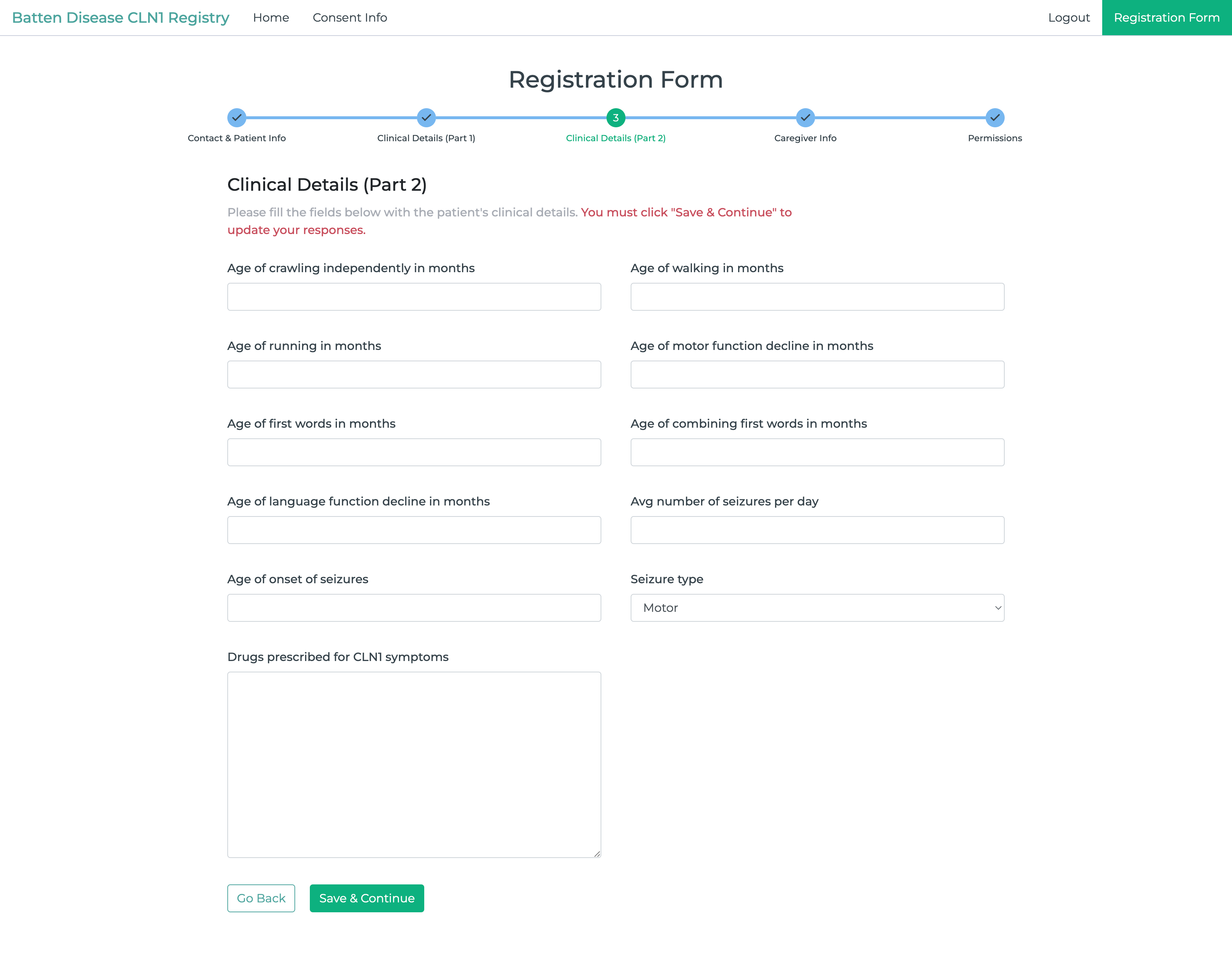

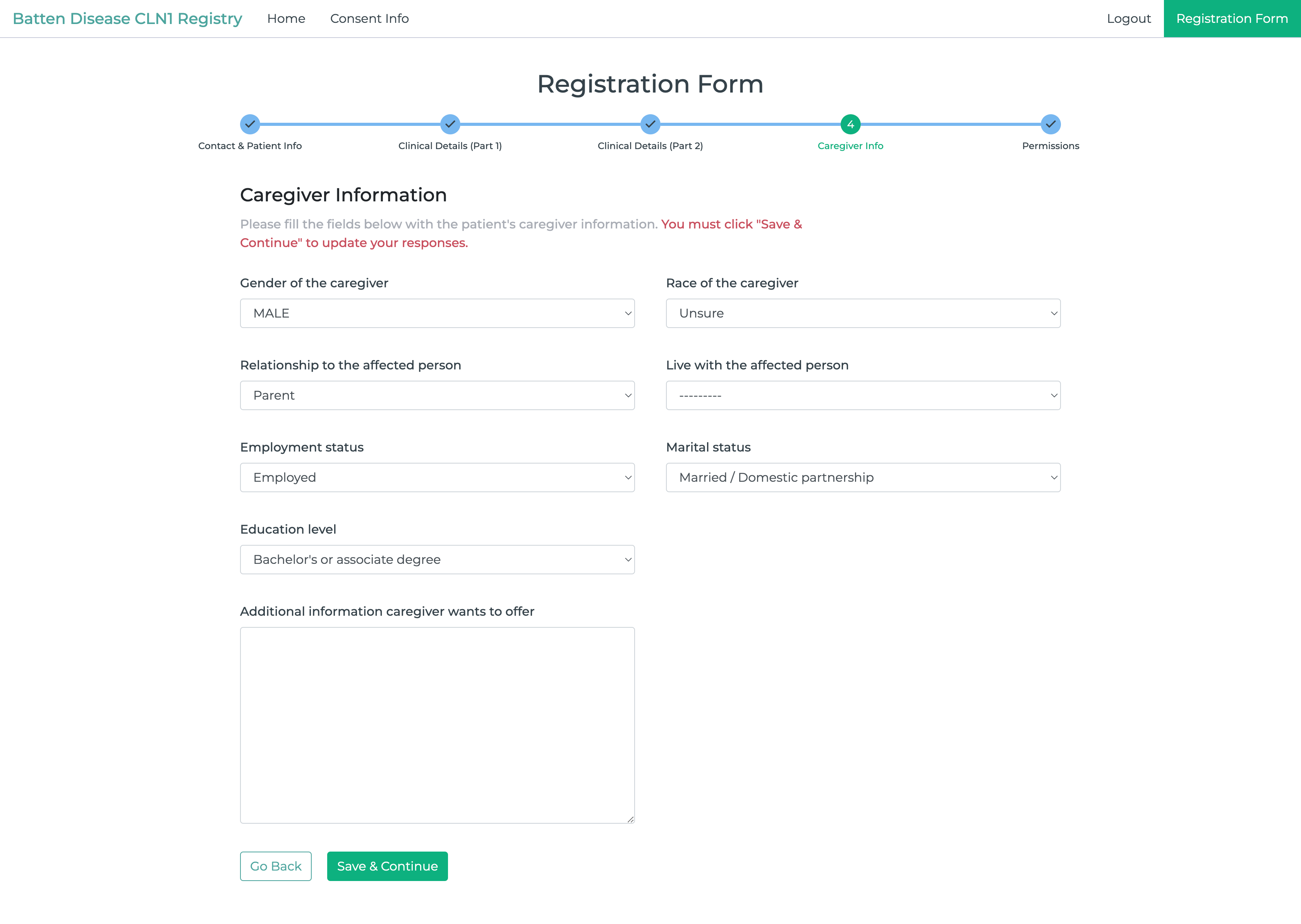

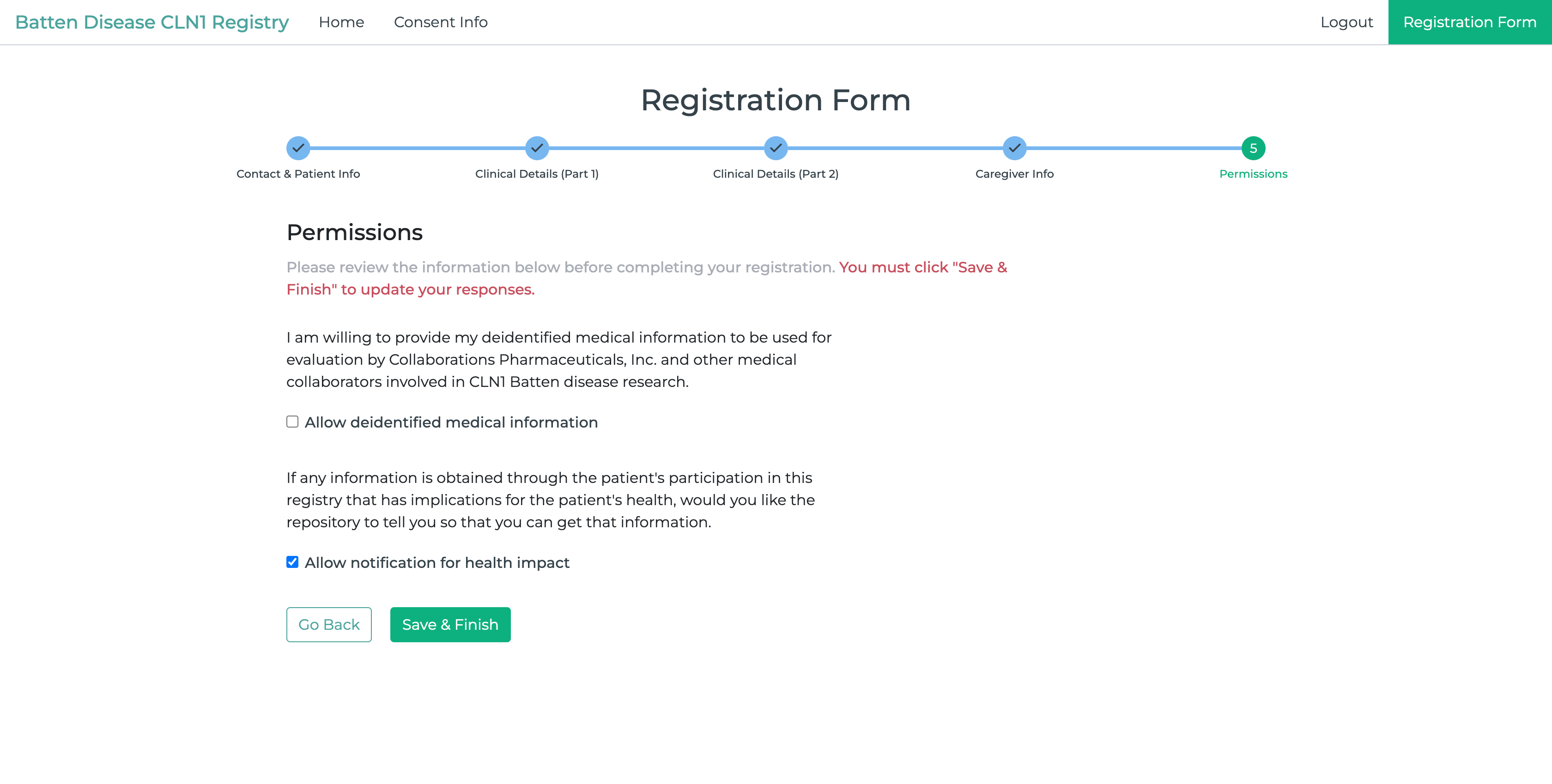

**Figure S2.** Distribution plots from the deidentified data analyzed from the CLN1 registry with smooth average curve fit of the data. A. Gender of patient, B. Race of the affected person. C. Age of diagnosis for affected person, D. Age of motor function decline, E. Age of language function decline, F. First Clinical Sign, G. Age of onset of seizures, H. Estimation in years of the affected persons mental age, I. Current age of affected person, J. Number of hospitalizations in the last year, K. number of episodes of pneumonia in the last year, L. Age of first words, M. Age of combining first words, N. Age of crawling independently, O. Age of walking, P. Age of running, Q. Number of siblings with CLN1. R. Number of siblings without CLN1, S. Average number of seizures per day, T. Seizure type, U. Time in years after noticing symptoms did CLN1 diagnosis take place, V. Drug classes used to treat symptoms in the CLN1 patient registry. W. Drugs used to treat symptoms in the CLN1 patient registry. X. First clinical sign grouped by disease classification.

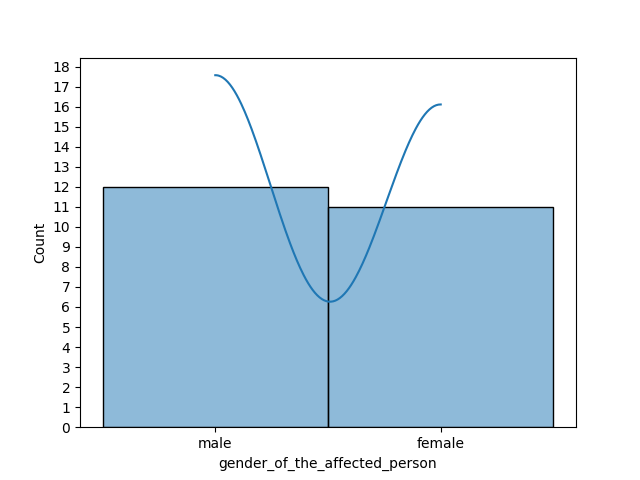

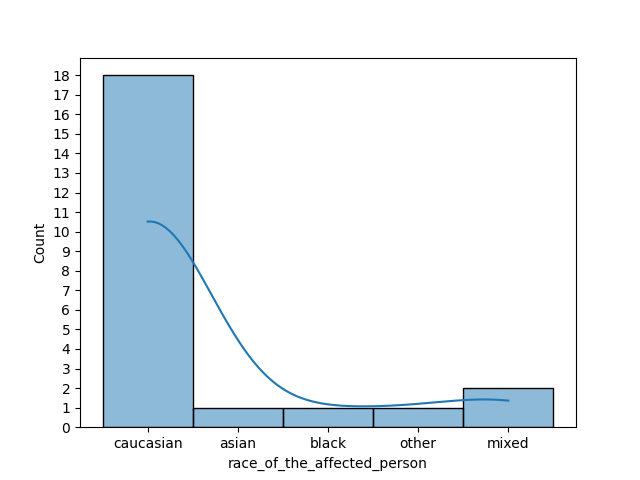

C

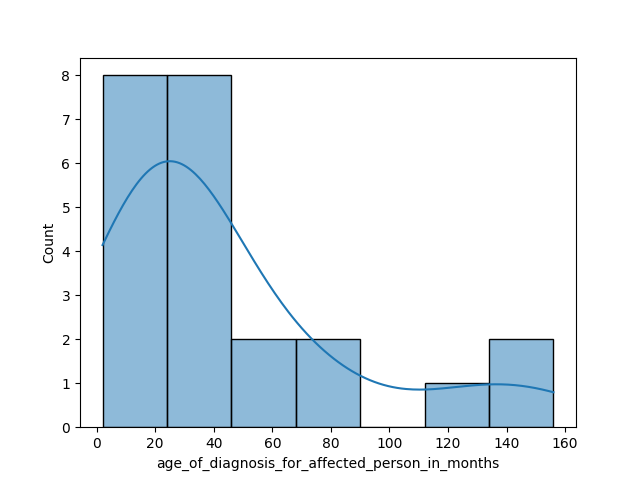

D

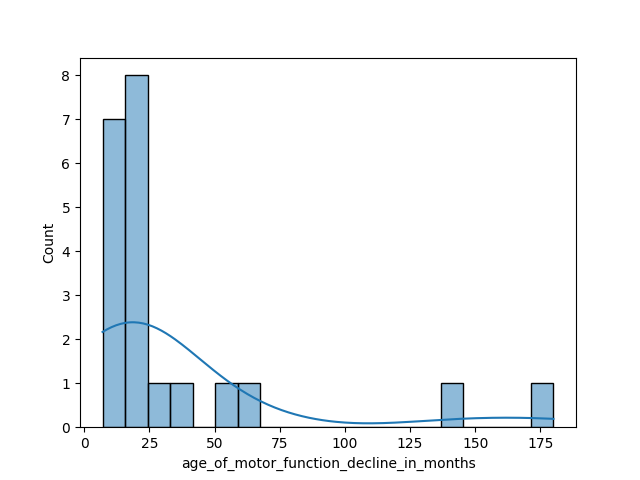

E

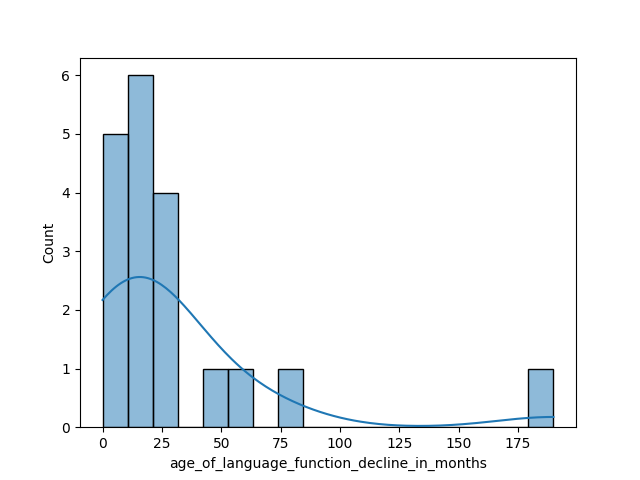

F

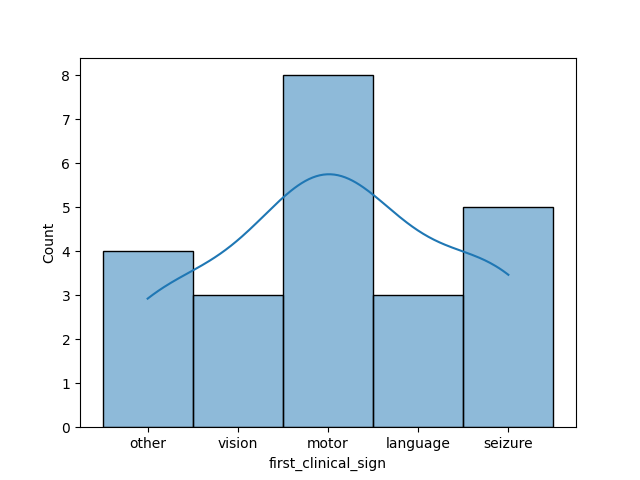

G

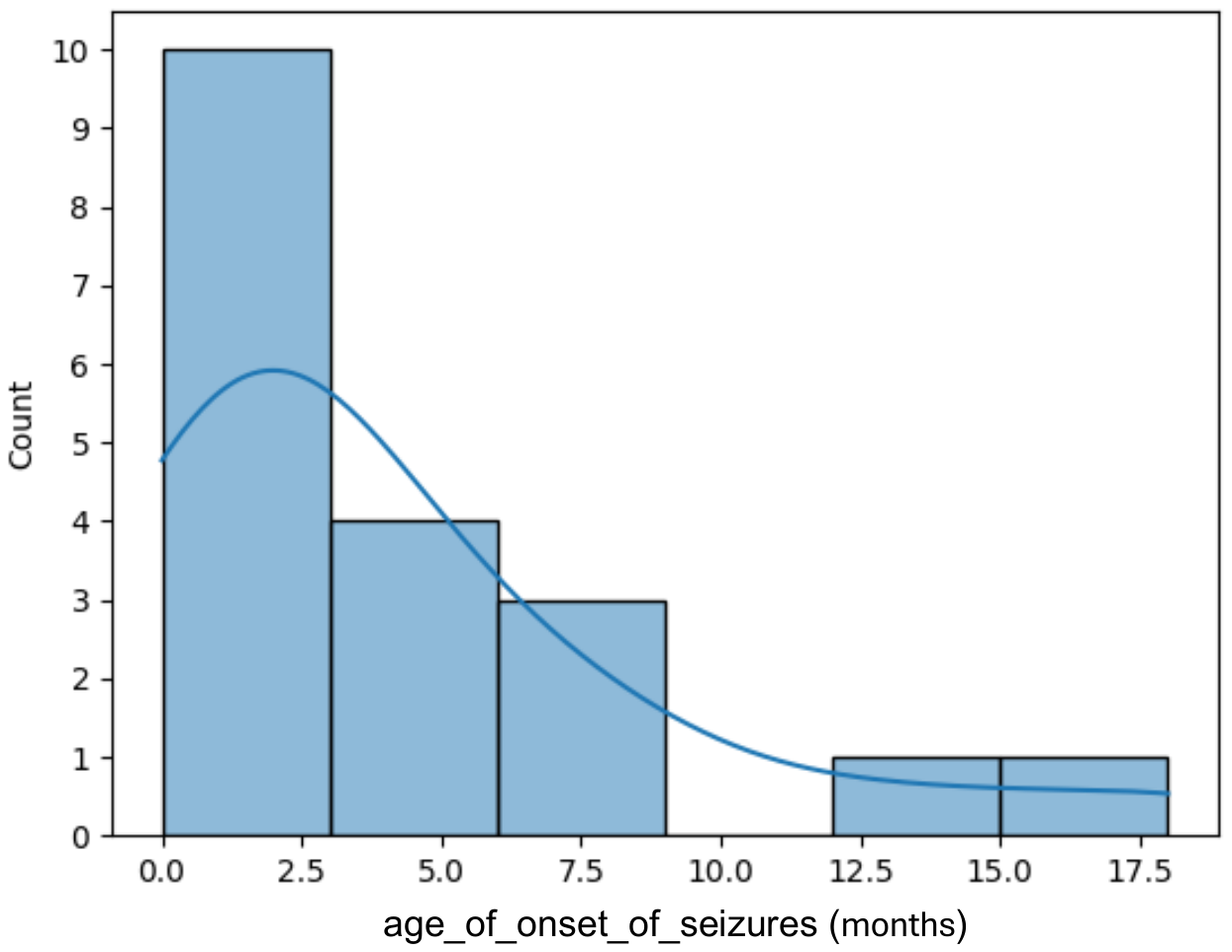

H

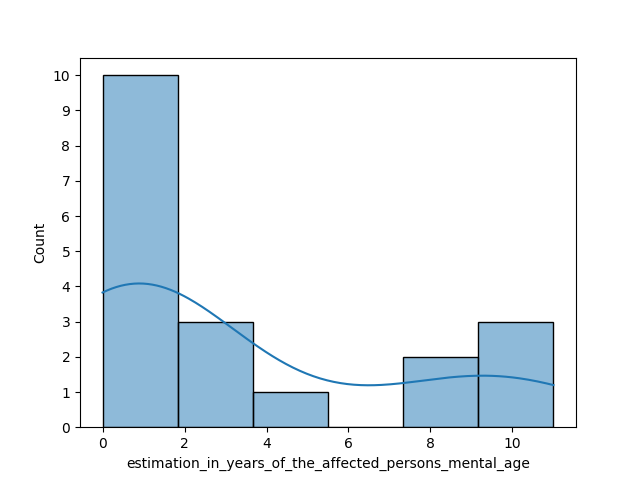

I

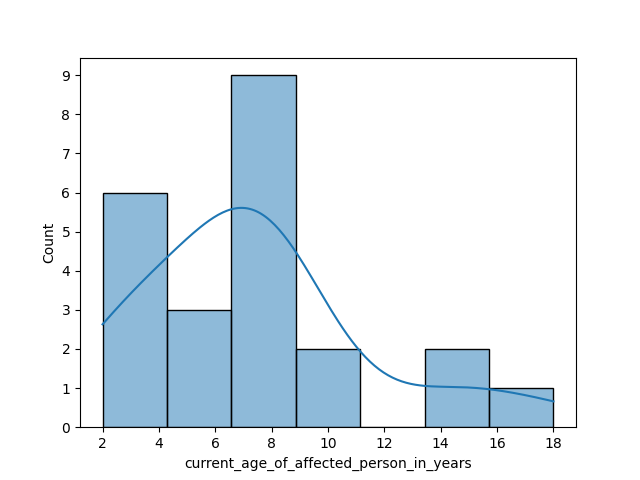

J

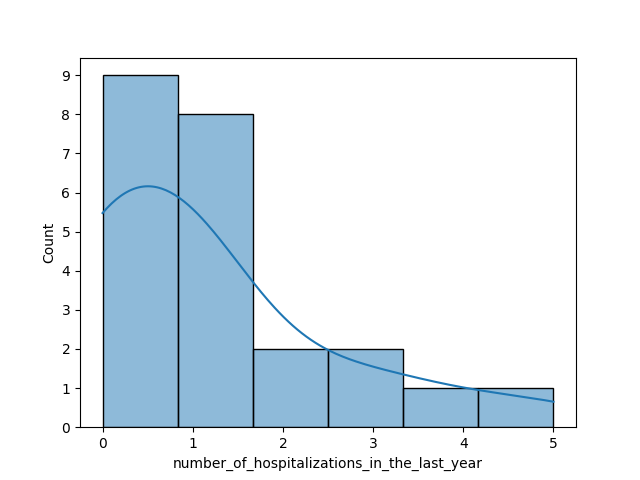

K

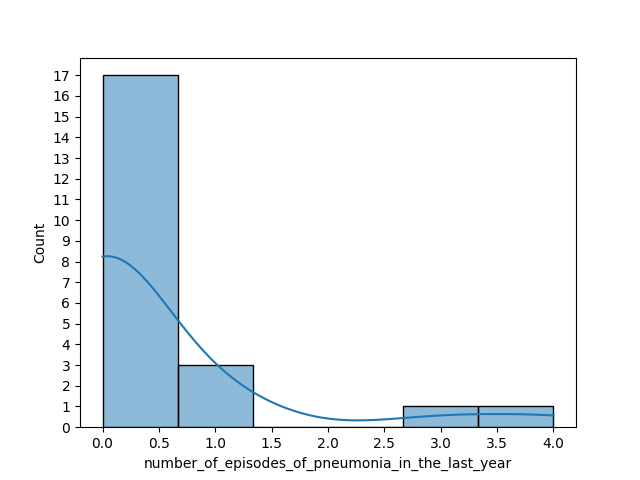

L

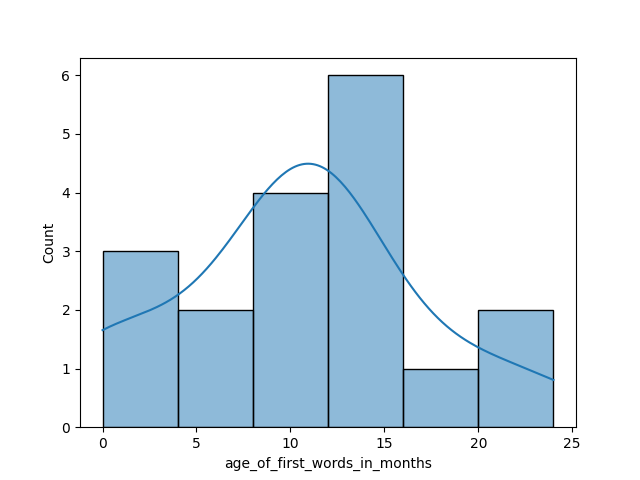

M

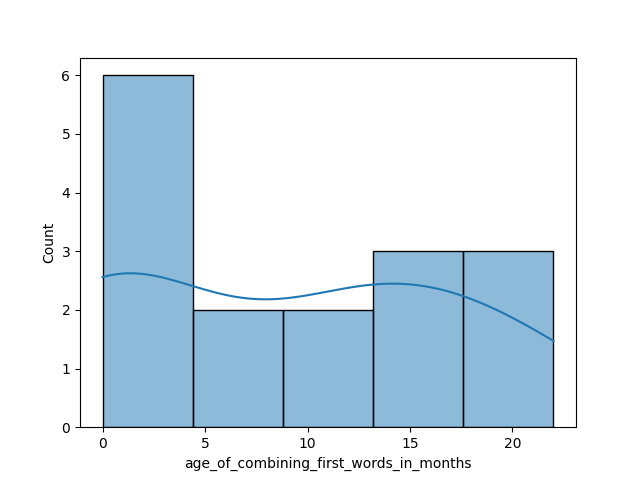

N

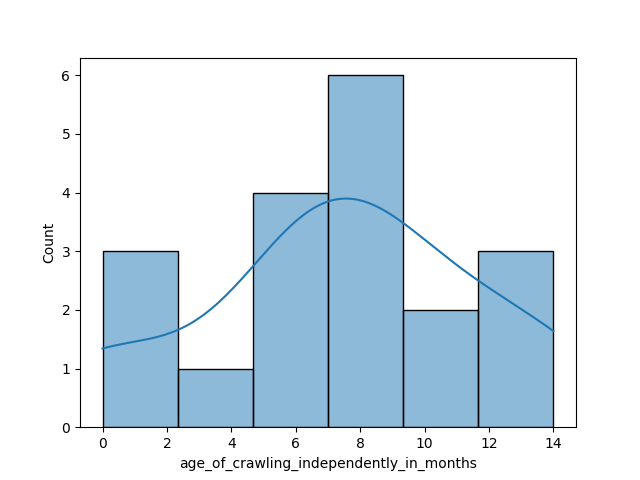

O

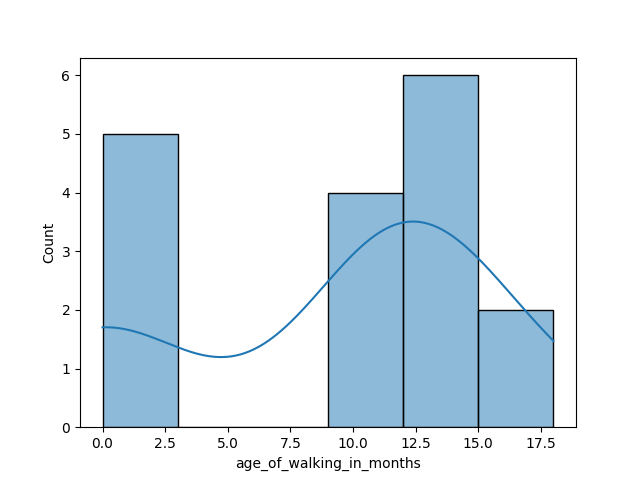

P

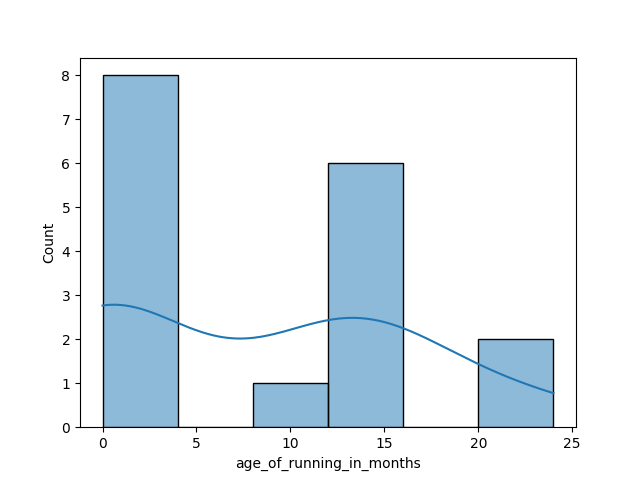

Q

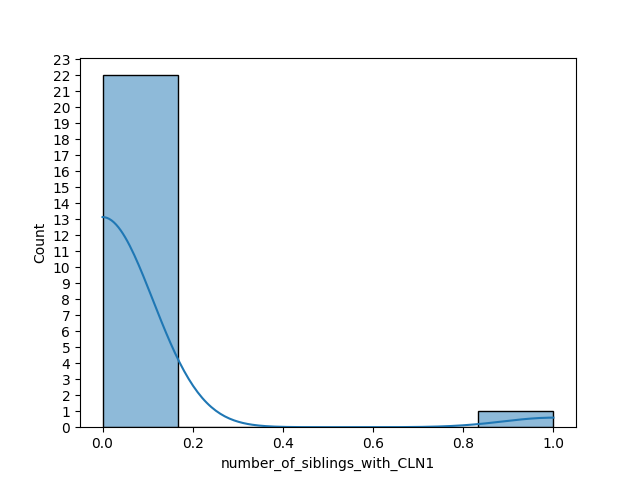

R

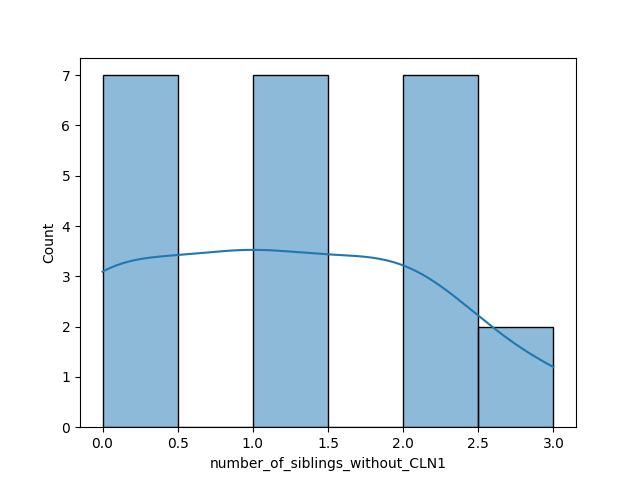

S

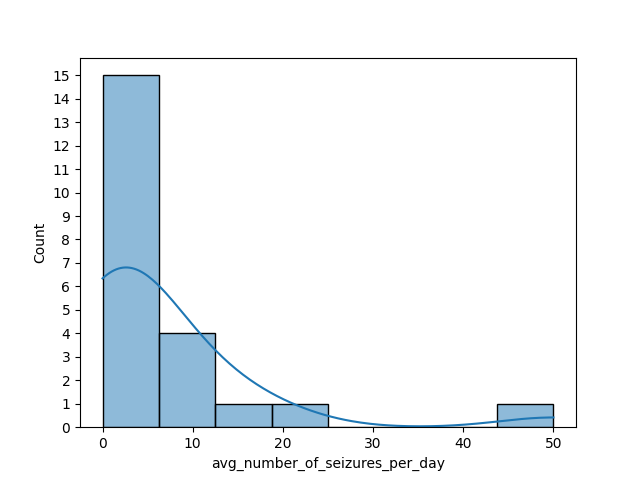

T

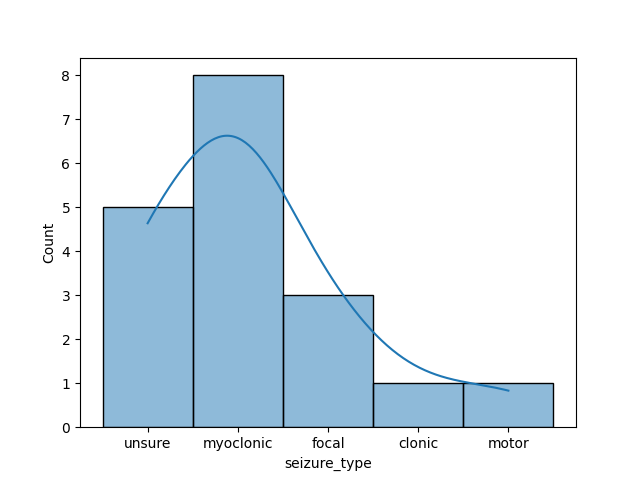

U

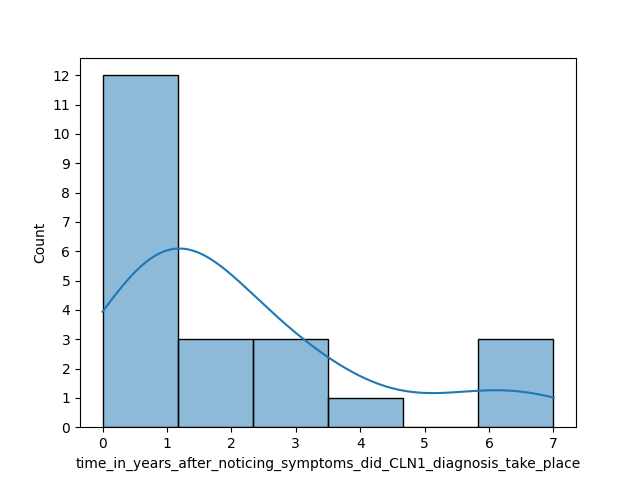

V

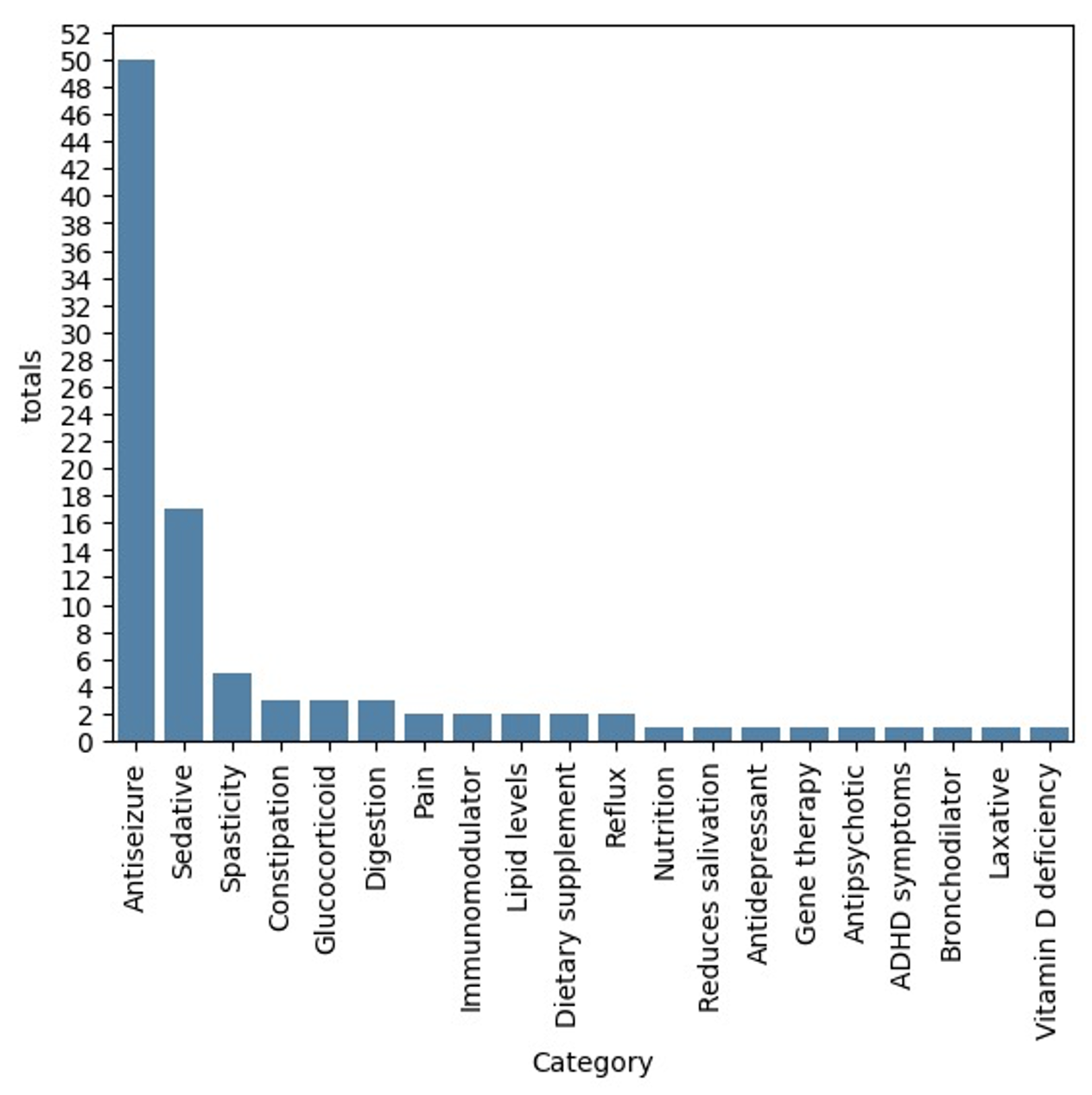

W

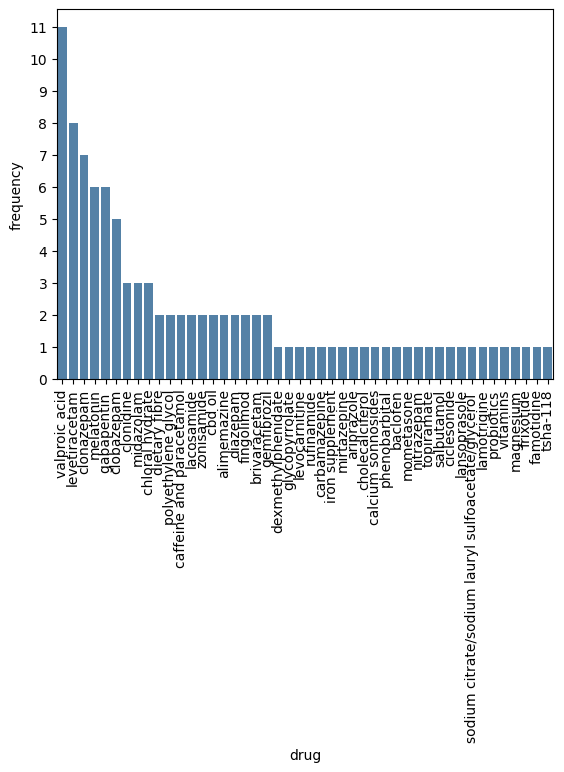

X

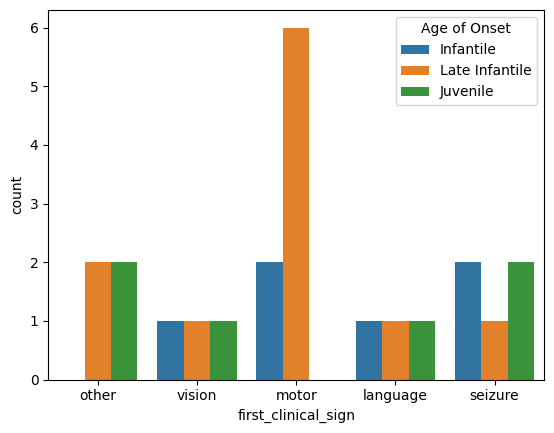

**Supplemental Table 1.** Summary data for questions from Batten Disease CLN1 Registry (Mean ± SD). ^#^ Incomplete data.

| **Class** | **N** | **Current age (Y)** | **Age of diagnosis (M)** | **Age of motor function decline (M)^#^** | **Age of language function decline (M)^#^** | **Ave number of seizures/ day^#^** | **Age of onset of seizures (Y)^#^** |
| --- | --- | --- | --- | --- | --- | --- | --- |
| Infantile (< 18 months) | 6 | 6.66 ± 2.16 | 11.67 ± 7.17 | 22.83 ± 18.99 | 28.25 ± 32.19 | 6.83 ± 6.31 | 2.60 ± 3.05 |
| Late Infantile (18 months - <4 yrs) | 11 | 5.54 ± 2.73 | 30.09± 8.94 | 16.63 ± 5.97 | 17.5 ± 4.90 | 9.27 ± 14.69 | 3.63 ± 4.99 |
| Juvenile (4 yr - < 18 yrs) | 6 | 11.33 ± 4.97 | 104.0 ± 39.77 | 104.0 ± 69.05 | 80.25 ± 74.50 | 0.8 ± 1.10 | 9 ± 3.61 |

**Supplemental Table 2.** Summary data for questions from Batten Disease CLN1 Registry (Mean ± SD). ^#^ Incomplete data.

| **Class** | **Time after noticing symptoms did CLN1 diagnosis take place (years)^#^** | **Estimation of the affected persons mental age (years)^#^** | **Number of hospitalizations in the last year** | **Number of episodes of pneumonia in the last year** | **Number of siblings without CLN1** | **Number of siblings with CLN1** |
| --- | --- | --- | --- | --- | --- | --- |
| Infantile (< 18 months) | 2.67 ± 2.58 | 3.60 ± 4.1 | 0.5 ± 0.55 | 0 ± 0 | 0.67 ± 0.82 | 0 ± 0 |
| Late Infantile (18 months - <4 yrs) | 1.91 ± 1.92 | 0.89 ± 1.27 | 1.82 ± 1.72 | 0.9 ± 1.45 | 1.27 ± 1.01 | 0 ± 0 |
| Juvenile (4 yr - < 18 yrs) | 2.2 ± 1.48 | 7.2 ± 4.32 | 0.67 ± 0.82 | 0.17 ± 0.41 | 1.5 ± 1.05 | 0.17 ± 0.41 |

**Supplemental Table 3.** Summary data for questions from Batten Disease CLN1 Registry (Mean ± SD). ^#^ Incomplete data.

| **Class** | **Age of crawling independently (months) ^#^** | **Age of walking (months) ^#^** | **Age of running (months) ^#^** | **Age of first words (months) ^#^** | **Age of combining first words (months) ^#^** |
| --- | --- | --- | --- | --- | --- |
| Infantile (< 18 months) | 9.0 ± 4.36 | 13.0 ± 2 | 15.33 ± 4.51 | 10.25 ± 3.50 | 10.0 ± 7.07 |
| Late Infantile (18 months - <4 yrs) | 9.17 ± 2.93 | 14.0 ± 2.83 | 14.5 ± 0.71 | 12.83 ± 5.60 | 13.0 ± 1.73 |
| Juvenile (4 yr - < 18 yrs) | 8.0 ± 2.12 | 11.6 ± 1.52 | 12.6 ± 7.80 | 13.4 ± 5.73 | 16.8 ± 5.76 |

**Supplemental Table 4.** Table of p-values to assess statistical significance in the Post-Hoc Dunn’s follow-up test. p-value significance <0.5-0.0332*, <0.0332-0.0021**, <0.0021-0.0002***, <0.0001****.

|  | Post-Hoc Dunn's Test for “Age of diagnosis for affected person in months” | | |
| --- | --- | --- | --- |
|  | Infantile | Juvenile | Late infantile |
| Infantile | 1 | 0.000041 | 0.026747 |
| Juvenile | 0.000041 | 1 | 0.026747 |
| Late infantile | 0.026747 | 0.026747 | 1 |
|  | Post-Hoc Dunn's Test for “estimation in years of the affected person’s mental age” | | |
|  | Infantile | Juvenile | Late infantile |
| Infantile | 1 | 0.208518 | 0.313435 |
| Juvenile | 0.208518 | 1 | 0.012866 |
| Late infantile | 0.313435 | 0.012866 | 1 |
|  | Post-Hoc Dunn's Test for “age of motor function decline in months” | | |
|  | Infantile | Juvenile | Late infantile |
| Infantile | 1 | 0.08404 | 0.927689 |
| Juvenile | 0.08404 | 1 | 0.048671 |
| Late infantile | 0.927689 | 0.048671 | 1 |
